# Nigrostriatal dopaminergic integrity in relation to prefrontal cortex activity and gait performance during dual-task walking in older adults

**DOI:** 10.1101/2025.02.28.25323092

**Authors:** Andrea L Rosso, Emma M Baillargeon, Nico I Bohnen, Brian J Lopresti, Theodore J Huppert, Lana M Chahine, Erin Jacobsen, Caterina Rosano

**Author notes:** **Corresponding author:** Andrea L Rosso.

## Abstract

Greater walking automaticity facilitates maintenance of gait speed without prefrontal cortex (PFC) resources. Brain aging may cause shifts to more attentional gait with greater engagement of the PFC. Nigrostriatal dopaminergic integrity likely facilitates walking automaticity and maintenance of gait speed during attentional dual-tasks. Older adults (n=201; age=74.9; 63.2% women, gait speed=1.10 m/s) completed a dual-task protocol of saying every other letter of the alphabet while walking. PFC activation was measured by functional near-infrared spectroscopy. Four groups were defined based on PFC activation (increased during dual-task, PFC+, or not, PFC-) and gait speed performance (maintained during dual-task, gait+, or slowed, gait-). To compare nigrostriatal dopaminergic integrity across groups, we assessed binding of the type-2 vesicular monoamine transporter (VMAT2) density in the sensorimotor and associative striatum using [^11^C]-(+)-α-dihydrotetrabenazine (DTBZ) positron emission tomography. Multinomial regression estimated adjusted associations of DTBZ binding with group membership. We hypothesized that DTBZ binding was highest for those who maintained gait speed without additional PFC activation when switching from single-to dual-task (PFC-/gait+; i.e., highest gait automaticity). In bivariate analyses, the PFC-/gait+ group had the highest DTBZ binding in the sensorimotor striatum (p=0.05); binding in the associative striatum was similar across groups (p=0.1).

Results were similar in adjusted regression analyses; DTBZ binding in sensorimotor striatum was associated with lower likelihood to be in the PFC-/gait-(OR=0.28; 95% CI: 0.08, 0.94) or the PFC+/gait+ (OR=0.29; 95% CI: 0.10, 0.84) groups compared to PFC-/gait+ reference group. These results provide support for dopaminergic involvement in sustaining gait automaticity at older ages.

## Introduction

Walking has traditionally been viewed as primarily modulated in the brain by the motor cortex and subcortical structures. A body of recent evidence has established the prefrontal cortex (PFC) as a critical component of gait control, particularly in older adults and during complex walking conditions^1,2^. In many adults, walking is an automatic motor task with little need for PFC control. Motor automaticity is characterized by lower frontal and increased posterior-subcortical engagement when performing overlearned motor tasks. However, aging-related neural declines in the integrity of motor-control regions lead to reduced walking automaticity and an increased reliance on PFC function during walking^3,4^. This increased reliance on PFC activity can be compensatory, preserving walking performance despite declines in the sensorimotor regions, but is inefficient, using PFC resources that may otherwise be needed for competing attentional tasks^2,3^ which can result in increased fall risk^5^. However, increased PFC activity can also be found in the presence of performance deterioration when it is interpreted as neural inefficiency and may represent noisy signaling in the PFC^6^. Therefore, interpretation of PFC activation during walking tasks in older adults is challenging in the absence of information pertaining to performance changes^7^ or other neural resources controlling movement.

Nigrostriatal sensorimotor dopaminergic integrity contributes to motor learning and automaticity^8^ which suggests that greater sensorimotor dopaminergic integrity leads to ability to maintain gait speed without additional PFC inputs under challenging conditions, such as dual-task walking. This is supported by prior research demonstrating that lower dopamine transporter (DAT) expression in striatum, as measured by [^11^C]β-CFT positron emission tomography (PET), is related to slower gait speed and greater likelihood of parkinsonian signs in adults without Parkinson’s disease (PD)^9,10^. Conversely, dopaminergic integrity of the nigrostriatal associative regions could support compensatory PFC activation facilitating maintenance of gait speed under challenging conditions due to this region’s strong connectivity to the PFC and role in modulating activity of the PFC. Further, executive control function, which is mediated via dopaminergic signaling in the associative regions of the striatum, is strongly related to gait speed and motor control in older adults, particularly during dual-task walking^1^. However, prior studies with measures of nigrostriatal dopaminergic integrity have focused on gait speed under unchallenged conditions and haven’t explored adaptation to walking conditions with increased attentional demands, such as dual-task walking. In addition, there are few studies using multimodal methods to assess both PFC function during walking tasks and aspects of brain health related to walking automaticity, and none currently determining the role of dopaminergic integrity by PET imaging.

We tested whether the level of nigrostriatal dopaminergic integrity measured via type-2 vesicular monoamine transporter (VMAT2) binding from dihydrotetrabenazine ([^11^C]DTBZ) PET imaging^11^ of the sensorimotor and associative striatum was associated with the PFC activation and gait speed response when moving from single-to dual-task walking. We hypothesized that greater dopaminergic integrity of the somatosensory network supports more automatic walking behaviors, requiring less PFC engagement. This would manifest as more adaptive responses to dual-task walking where gait speed is maintained in the absence of greater activity of the PFC. We further hypothesized that greater dopaminergic integrity of the associative network would support greater engagement of the PFC as needed to maintain gait speed when switching from single-to dual-task gait speed. Therefore, we hypothesized the greatest DTBZ binding for sensorimotor striatum would be seen in the group that maintained gait speed from single-to dual-task with no additional activity in the PFC and the greatest DTBZ binding for the associative striatum would be in the group that maintained gait speed from single-to dual-task with additional PFC activity. These results will improve understanding of automatic motor control during walking and it’s decline during aging with implications for development of intervention development and future research.

## Methods

### Participants

Participants came from two studies with harmonized protocols for magnetic resonance (MR), PET and functional near infrared spectroscopy (fNIRS) data collection. A subset of participants from the Monongahela-Youghiogheny Healthy Aging Team (MYHAT) study were included in Move MYHAT (MMH), a substudy of dopaminergic contributions to mobility resilience. The parent study is a community-based study of risk factors for mild cognitive impairment. Recruitment of community-dwelling adults aged 65 and older in small towns in Southwest Pennsylvania occurred between 2006-2008 and 2016-2019 (n=2685), and annual follow-up continues^12,13^. Of 386 eligible and referred, MMH recruited participants (n=87) who were still in the study in 2019-2021 and could walk unassisted, were eligible for and agreed to participate in the MR and PET protocols, and were free of dementia or other neurodegenerative conditions, including PD.

The second study was an ancillary of the Study of Muscle, Mobility and Aging (SOMMA; n=879)^14^ to assess dopaminergic contributions to fatigability in older adults. SOMMA recruited at the University of Pittsburgh and Wake Forest University School of Medicine between April 2019 and December 2021. Individuals were eligible if they were aged 70 and older with gait speeds ≥0.6 m/s, who were able to complete a 400 m walk, were free of neurological diagnoses including PD and dementia, had a BMI<40 kg/m^2^, and were eligible for MRI and muscle biopsy. The SOMMA Brain ancillary further recruited individuals from the 439 participants enrolled at the Pittsburgh site between 2020-2022. Of the 285 potentially eligible participants whose muscle biopsy for the parent study was less than 12 months prior, a total sample of n=150 was recruited for the ancillary and completed neuroimaging. Data for SOMMA came from the November 2024 data release. Participants from both studies (n=85 from MMH and n=116 from SOMMA) were included in these analyses if they had usable data on all relevant measures.

All neuroimaging data (fNIRS, PET) was collected on the same instruments and the dual-task walking protocols were completed in the same lab space with the same data collection team for both studies. All protocols were approved by participating Institutional Review Boards and all participants gave informed consent prior to any data collection.

### Dual-task Walking

Participants walked a 15-meter straight path at their comfortable pace with and without a concurrent cognitive task (single- and dual-task walking)^15^. Single- and dual-task walking were each repeated four times and order of presentation was randomized. The dual-task consisted of reciting every other letter of the alphabet starting with ‘B’ while walking^16^. The alphabet task was also completed for 20 seconds while standing. Twenty second quiet standing periods were included before each task as a baseline condition for the fNIRS signal. All tasks were completed while wearing a wireless fNIRS system on the forehead (Artinis Octamon). Gait speed was measured by stopwatch and calculated in meters/second (m/s) for each walking task. Change in gait speed from single-to dual-task was calculated with maintenance of gait speed (i.e., less slowing) from single-to dual-task considered to be the healthier response. Rate of correct letters generated was calculated as the number of correct letters/s.

### fNIRS data collection and processing

We followed consensus recommendations in our fNIRS data processing and reporting^17^. Participants wore an eight-channel continuous wave fNIRS system across the forehead to measure changes in activity in the right and left PFC regions. The fNIRS instrument measured oxygenated hemoglobin (HbO) and deoxygenated hemoglobin (Hbr) concentrations at 850 and 760 nm, respectively, with one detector and four sources on each side of the head. The probe approximately covered bilateral Brodmann areas BA9, BA44, BA45, and BA46^18^. Optical data were collected at 10 Hz and stored using OxySoft software (Artinis Medical Systems).

Raw signals were exported to Matlab (MATLAB and Statistics Toolbox Release 2021b) for processing with the NIRS Brain AnalyzIR Toolbox^19^. Flat channels due to saturation or equipment error were removed from analysis. Visual checks were performed to confirm no time overlap across tasks and for data quality. Observations were excluded where protocol deviations (e.g., participant walked during a standing task) occurred. Because all tasks were completed four times, exclusion of some observations did not impact overall data availability or quality.

The fNIRS light intensity signals were converted to optical density and resampled to 4 Hz to reduce computational burden. The modified Beer-Lambert law was applied to generate concentration of HbO (in µM) from optical density data.^20^ In first-level modeling, a canonical model with autoregressive pre-whitening approach using iteratively reweighted least-squares (AR-IRLS) was implemented to model changes in HbO for each task relative to a global baseline.^21^ Previous work demonstrated that this iterative regression model, which uses a robust statistical estimator to downweigh outlier data points, is less prone to bias from motion artifacts in the fNIRS data, which result from slippage of the head cap.^21^ Channel-wise student’s t tests compared the level of HbO change for each task from the quiet standing baseline state. These coefficients from the linear regression were then used to combine the results of the four trials using a mixed-effects model with fixed effect of task. The mixed effects model used robust weighted least-square regression with the estimate noise covariance matrix derived from the first-level regression model. The channel-specific t-statistic values were averaged over the combined right and left hemispheres of the PFC for all subsequent analyses.

### PET Imaging

[^11^C]-(+)-a-DTBZ was synthesized using a modification of the methodology reported by Kilbourn, et al.^22^ with high molar activity (4.14 mCi/nmol at end-of-synthesis). The major modifications included the use of [^11^C]methyl triflate as the labelling synthon and the purification of the desired [^11^C]DTBZ using a semi-preparative HPLC column. Radiochemical and chemical purities were >95% as determined by analytical HPLC.

Simultaneous acquisition of [^11^C]DTBZ PET and 3 Tesla magnetic resonance (MR) images was performed on a Siemens Biograph mMR scanner (Siemens Medical Solutions USA, Malvern, PA)^23^. PET emission data were collected in list-mode over 60 min post-injection and were reconstructed using filtered back projection (FBP) with Fourier rebinning into a dynamic series of 20 frames ranging in duration from 15 sec to 300 sec. Attenuation correction of PET emission data was performed analytically using previously described model-based methods^24,25^. The [^11^C]DTBZ distribution volume ratio (DVR) was estimated from indices of [^11^C]DTBZ binding derived using the non-invasive Logan graphical analysis method^26^ using occipital cortex as a reference tissue. Volumes of interest (VOIs) for sampling of PET image data were defined on the MPRAGE MR series using an automated atlas-based method (CIC Atlas, Clinical Imaging Center, Imperial College, London, UK)^27^.

We included two *a priori* defined regions of interest: the sensorimotor striatum (posterior putamen) and the associative striatum (anterior putamen, predorsal caudate, postdorsal caudate). The sensorimotor striatum is primarily connected to sensorimotor regions of the cortex, including the primary and supplementary motor cortices, which are involved in automatic motor control. The associative striatum is involved in executive functions and has strong connections to the associative cortex, including the PFC. VMAT2 was chosen as the target as it is responsible for the translocation of monoamine neurotransmitters from the cytoplasm into synaptic vesicles. The use of VMAT2 as an index of nigrostriatal dopaminergic integrity is predicated on the fact that ∼95% of monoaminergic synapses in the striatum are dopaminergic^28,29^.

### Covariates

Age at MRI, sex, race, and education were self-reported. Due to low numbers of non-White participants, we categorized race as either White or non-White which included 17 Black, 1 Asian, and 1 Native American participants, and 1 participant with unreported race. Education was recorded as completed less than high school, high school or GED, or greater than high school; this was recoded as less than or completed high school compared to those with more than high school education. Cognitive status was harmonized across the two datasets as either unimpaired or impaired. In MMH, cognitive impairment was determined in the parent study by Clinical Dementia Rating (CDR)^30^ of 0.5. In SOMMA, cognitive impairment was derived from age, sex, race, and education adjusted percentile scores on the Montreal Cognitive Assessment (MoCA)^31^ in the SPRINT trial^32^; the 10^th^ percentile was used to identify both mild cognitive impairment and dementia. Body mass index (BMI) was calculated from measured height and weight. Systolic blood pressure was recorded by a single measurement in MMH and by the average of two measurements in SOMMA. Health conditions in each study are coded as presence of any of self-reported cancer (excluding nonmelanoma skin cancer), cardiac arrhythmia, chronic kidney disease, chronic obstructive pulmonary disease (COPD), coronary artery disease (myocardial infarction), congestive heart failure, diabetes, and stroke. Structural brain measures were obtained by structural 3T MR imaging concurrent with PET imaging. 3T MR images were acquired by T1-weighted magnetization-prepared rapid gradient echo (MPRAGE) and Dixon sequences^33^. Total white matter hyperintensities were assessed using the lesion segmentation tool^34^ for the MRI analysis toolbox SPM-12. White matter hyperintensity volumes were normalized to the total cerebral white matter volume. Brain atrophy was defined as the ratio of total grey matter volume to intracranial volume.

### Statistical Analysis

All analyses were conducted in SAS 9.4 using the combined dataset for the two studies. Variables are presented as either means with standard deviations (SD) or as n and percent for categorical variables for the combined sample and for each study individually.

We defined four groups based on PFC and gait speed change when transitioning from single- to dual-task walking (Figure 1). PFC change was defined by the t-statistic from the fNIRS signal as either a negative or 0 change (PFC-) or a positive change (PFC+) from single-to dual-task. Gait speed change was defined using a threshold of 0.1 m/s decline; those having less than this change were considered to maintain speed from single-to dual-task (gait+) and those with more than 0.1 m/s decline were considered to have slowed (gait-). Four groups were then generated: 1) PFC-/gait+ who did not have an increase in PFC activation and maintained or increased gait speed from single-to dual-task walking, 2) PFC-/gait-who did not have an increase in PFC activation and had a decline in gait speed from single-to dual-task walking, 3) PFC+/gait+ who had an increase in PFC activation and maintained or increased gait speed from single-to dual-task walking, and 4) PFC+/gait-who had an increase in PFC activation with a concurrent decline in gait speed from single-to dual-task walking.

**Figure 1.**
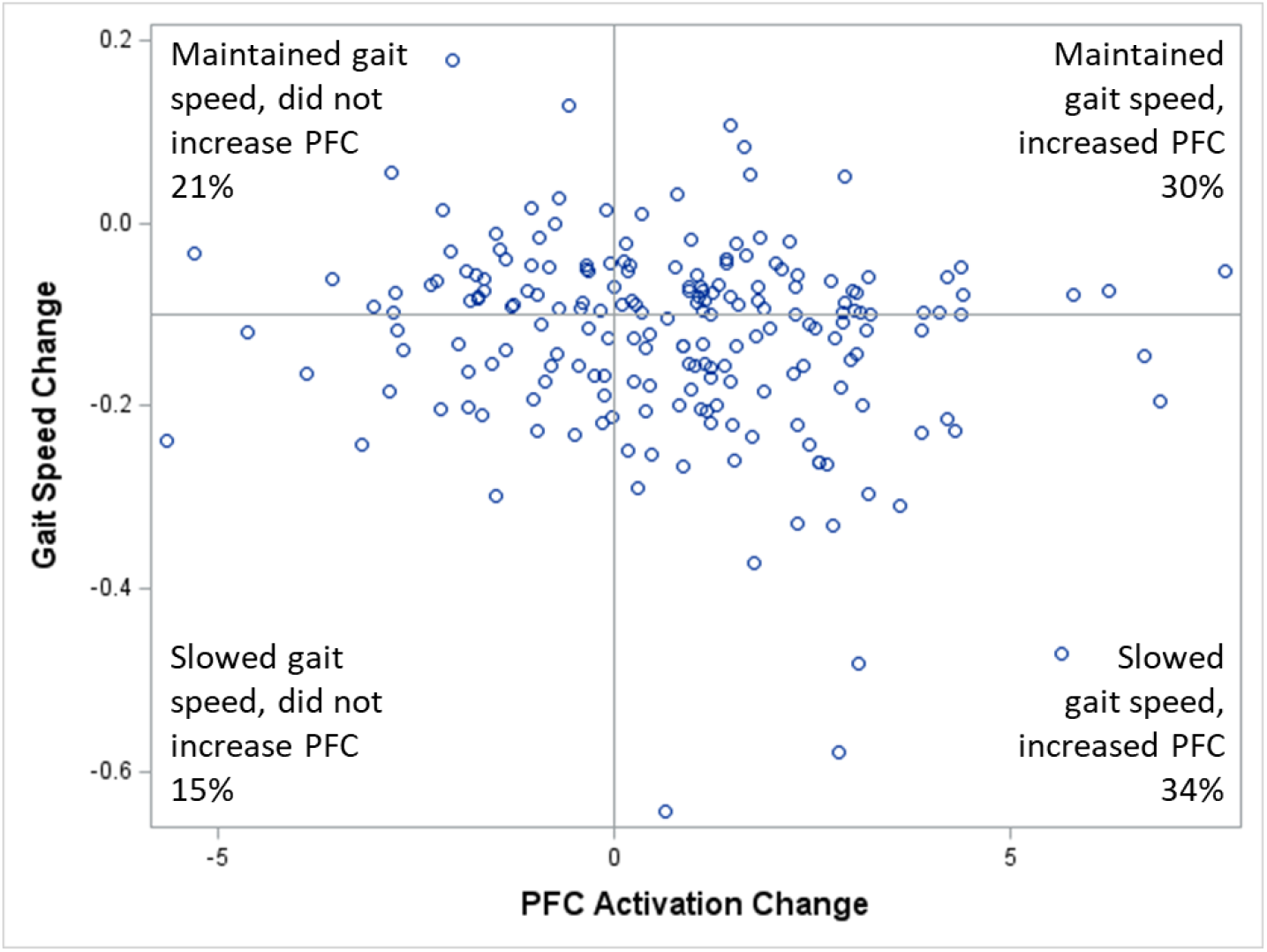
Change from single-to dual-task walking in prefrontal cortex (PFC) activation by functional near-infrared spectroscopy and gait speed (m/s) in older adults (n=201). Analytic groupings are shown along with prevalence of each.

The relation of demographic, health, performance and neuroimaging variables with the PFC/gait groups was assessed by Chi-square tests for categorical variables, except for race and cognitive impairment where Fisher’s exact test was used to account for small sample sizes within cells. Continuous variables were compared across PFC/gait groups using ANOVA. Multinomial logistic regression was used to assess the relation of DTBZ binding levels as the independent variable with the PFC/gait groups as the dependent variable with adjustment for covariates. Group 1 (PFC-/gait+) served as the reference group for all analyses. Separate models were built with DTBZ in either the sensorimotor striatum or in the associative striatum as the independent variable. Due to the limited sample size, covariates of study, age, sex, race, education, BMI, blood pressure, presence of health conditions, WMH and brain atrophy were assessed in the model one at a time. Covariates that were significant at p<0.05 in these models were then retained in the final adjusted model. While study, age and sex were not significant in the individual models, they were felt to be important confounders and were retained in the final adjusted model, along with BMI and the brain atrophy index which were significantly associated. Sensitivity analyses assessed whether gait performance alone or PFC activation alone was related to DTBZ binding and also excluded those with cognitive impairment.

## Results

The analytic sample combined across the two cohorts (n=201, Table 1) was 75 years old on average and predominantly women (63%), White (92%) and with a greater than high school education (83%). In the total sample, 50% had at least one health condition and 7% were cognitively impaired. Mean single-task gait speed was 1.10 (SD=0.20) m/s and mean change in gait speed from single-to dual-task was -0.13 (SD=0.12) m/s.

**Table 1.** Demographic, health, neuroimaging, and dual task walking characteristics by group based on prefrontal cortex (PFC) activation and gait performance from single-to dual-task walking in older adults.

|  |  | Did not increase PFC (PFC-) |  | Increased PFC (PFC+) |  |  |
| --- | --- | --- | --- | --- | --- | --- |
|  | Total | maintained gait (gait+) | slowed down (gait-) | maintained gait (gait+) | slowed down (gait-) |  |
|  | N=201 | N=42, 21% | N=30, 15% | N=61, 30% | N=68, 34% |  |
|  | Mean (SD) or n (%) | Mean (SD) or n (%) | Mean (SD) or n (%) | Mean (SD) or n (%) | Mean (SD) or n (%) | p-value |
| Age (years) | 74.9 (4.6) | 74.1 (4.2) | 74.4 (4.7) | 75.1 (4.6) | 75.3 (4.8) | 0.5 |
| Women | 127 (63.2%) | 26 (61.9%) | 19 (63.3%) | 38 (62.3%) | 44 (64.7%) | 0.9 |
| White race | 185 (92.0%) | 40 (95.2%) | 27 (90.0%) | 56 (91.8%) | 62 (91.2%) | 0.9* |
| > HS education | 165 (82.5%) | 32 (76.2%) | 26 (86.7%) | 52 (86.7%) | 55 (80.9%) | 0.5 |
| BMI | 28.4 (5.5) | 30.3 (6.2) | 29.1 (5.7) | 28.5 (4.7) | 26.9 (5.3) | 0.02 |
| Systolic blood pressure (mmHg; n=173) | 130.8 (15.0) | 131.0 (13.3) | 134.9 (15.1) | 130.5 (15.7) | 128.8 (15.4) | 0.4 |
| Any health condition | 100 (50.0%) | 22 (53.7%) | 13 (43.3%) | 27 (44.3%) | 38 (55.9) | 0.5 |
| Cognitively unimpaired | 186 (93.0%) | 39 (92.9%) | 27 (90.0%) | 57 (95.0%) | 63 (92.7%) | 0.8* |
| Study (SOMMA) | 116 (57.7%) | 21 (50.0%) | 16 (53.3%) | 39 (63.9%) | 40 (58.8%) | 0.5 |
| WMH (mm<sup>2</sup>; n=197) | 0.014 (0.02) | 0.021 (0.046) | 0.011 (0.010) | 0.014 (0.012) | 0.012 (0.018) | 0.3 |
| Brain atrophy | 0.34 (0.03) | 0.34 (0.03) | 0.34 (0.03) | 0.33 (0.03) | 0.33 (0.03) | 0.08 |
| *Dual-Task* |  |  |  |  |  |  |
| HbO change single- to dual-task (t-statistic)** | 0.78 (2.21) | -1.45 (1.08) | -1.56 (1.42) | 2.02 (1.55) | 2.08 (1.45) | <0.001 |
| Gait speed change single- to dual-task (m/s)** | -0.13 (0.12) | -0.04 (0.06) | -0.18 (0.05) | -0.06 (0.04) | -0.22 (0.14) | <0.001 |
| Gait speed, single task (m/s) | 1.10 (0.20) | 1.08 (0.21) | 1.07 (0.20) | 1.09 (0.16) | 1.14 (0.23) | 0.3 |
| Rate of correct letters, dual task (letters/s) | 0.60 (0.19) | 0.63 (0.19) | 0.57 (0.13) | 0.61 (0.21) | 0.57 (0.17) | 0.3 |
| *DTBZ binding* |  |  |  |  |  |  |
| Sensorimotor striatum | 2.44 (0.43) | 2.58 (0.48) | 2.38 (0.45) | 2.35 (0.42) | 2.46 (0.38) | 0.05 |
| Associative Striatum | 2.09 (0.34) | 2.17 (0.39) | 2.05 (0.40) | 2.02 (0.33) | 2.12 (0.28) | 0.1 |

\*Fisher's exact test
\*\* These variables only differ across levels that define the groups
HS=high school, BMI=body mass index, WMH=white matter hyperintensities, DTBZ= dihydrotetrabenazine, HbO=oxygenated hemoglobin, m/s=meters/second

There were few notable differences across the two cohorts, but a greater proportion of participants in SOMMA had a greater than high school education (92%) compared with MMH (69%). Mean values and standard deviations for neuroimaging variables (MRI, PET, and fNIRS) were comparable across the two cohorts (Supplementary Table 1). Performance during dual-task walking for both gait speed and rate of correct letters generated was also highly comparable across the cohorts (Supplementary Table 1).

The sample was fairly evenly split among the four PFC/gait groups, though PFC+/gait-was the most common (34% of the sample) whereas PFC-/gait-was the least common (15% of the sample; Figure 1). Most demographic and health characteristics did not differ by PFC/gait group (Table 1). BMI was highest for the PFC-/gait+ group (mean=30.3) and lowest for the PFC+/gait- group (mean=26.9, p=0.02). Change in HbO signal and gait speed from single-to dual-task significantly differed by group (p<0.001 for each) as these variables were used to define the groups. Gait speed during single-task walking and rate of correct letters generated on the cognitive task alone did not differ among groups (p=0.3 for each). DTBZ binding in the sensorimotor striatum was significantly different between PFC/gait groups in bivariate analyses; PFC-/gait+ had the highest levels of DTBZ binding (mean=2.58) whereas PFC+/gait+ had the lowest (mean= 2.35; p=0.05). There were no significant differences in DTBZ binding in the associative striatum across groups in bivariate analyses (p=0.1).

In adjusted multinomial regression analyses (Table 3), those with greater DTBZ binding in the sensorimotor striatum were less likely to be in the PFC-/gait- (OR=0.28; 95% CI: 0.08, 0.95) or the PFC+/gait+ (OR=0.29; 95% CI: 0.10, 0.84) groups compared to the PFC-/gait+ reference group. There was no difference in DTBZ binding in the sensorimotor striatum for those in the PFC+/gait-compared to the PFC-/gait+ reference group. In adjusted multinomial regression analyses for DTBZ binding in the associative striatum, there was a significant difference for those in the PFC+/gait+ group compared to the PFC-/gait+ reference group (OR=0.23; 95% CI: 0.06, 0.85). No other significant differences were observed for DTBZ binding in the associative striatum although ORs were comparable to those for the sensorimotor striatum.

In sensitivity analyses that defined groups based solely on PFC activation or on gait performance (rather than on the combination of the two), there were no associations of DTBZ binding with either PFC activation alone or gait performance alone (Table 3). In additional sensitivity analyses excluding those with cognitive impairment (Table 2), results were largely unchanged with the exception that the association of DTBZ binding at the sensorimotor striatum for PFC-/gait-compared to PFC-/gait+ was no longer significant.

**Table 2.** Adjusted multinomial logistic regression models of dihydrotetrabenazine (DTBZ) binding for dopaminergic neurotransmission by region of interest with groups based on prefrontal cortex (PFC) activation and gait performance from single-to dual-task walking in older adults (n=201).

|  | Did not increase PFC (PFC-) |  | Increased PFC (PFC+) |  |
| --- | --- | --- | --- | --- |
|  | maintain gait<br>(gait+) | slowed down<br>(gait-) | maintained gait<br>(gait+) | slowed down<br>(gait-) |
|  | N=42 | N=30 | N=61 | N=68 |
|  | OR (95% CI) | OR (95% CI) | OR (95% CI) | OR (95% CI) |
| **Sensorimotor striatum** |  |  |  |  |
| Unadjusted | REF | **0.31 (0.10, 0.97)** | **0.28 (0.10, 0.76)** | 0.48 (0.18, 1.24) |
| Adjusted* | REF | **0.28 (0.08, 0.95)** | **0.29 (0.10, 0.84)** | 0.53 (0.18, 1.54) |
| Adjusted* CN only** | REF | 0.33 (0.09, 1.18) | **0.27 (0.09, 0.83)** | 0.47 (0.15, 1.44) |
| **Associative Striatum** |  |  |  |  |
| Unadjusted | REF | 0.34 (0.08, 1.41) | **0.27 (0.08, 0.92)** | 0.63 (0.20, 2.04) |
| Adjusted* | REF | 0.27 (0.06, 1.23) | **0.23 (0.06, 0.85)** | 0.49 (0.13, 1.82) |
| Adjusted* CN only** | REF | 0.34 (0.07, 1.63) | **0.21 (0.05, 0.80)** | 0.47 (0.12, 1.82) |

\*Adjusted for study, age, sex, body mass index, and brain atrophy
\*\*n=186
CI=confidence interval, CN=cognitively normal, OR=odds ratio, REF=reference

**Table 3.** Adjusted logistic regression models of dihydrotetrabenazine (DTBZ) binding for dopaminergic neurotransmission by region of interest with groups based on either prefrontal cortex (PFC) activation or change in gait performance from single-to dual-task walking in older adults (n=201).

|  | Did not increase<br>PFC (PFC-) | Increased PFC<br>(PFC+) | maintained gait<br>(gait+) | slowed down<br>(gait-) |
| --- | --- | --- | --- | --- |
|  | N=72 | N=129 | N=103 | N=98 |
|  | OR (95% CI) | OR (95% CI) | OR (95% CI) | OR (95% CI) |
| **Sensorimotor striatum** |  |  |  |  |
| Unadjusted | REF | 0.63 (0.31, 1.25) | REF | 0.90 (0.47, 1.72) |
| Adjusted* | REF | 0.70 (0.33, 1.49) | REF | 0.92 (0.45, 1.87) |
| Adjusted* CN<br>only** | REF | 0.59 (0.27, 1.30) | REF | 0.94 (0.45, 1.97) |
| **Associative Striatum** |  |  |  |  |
| Unadjusted | REF | 0.67 (0.29, 1.60) | REF | 1.13 (0.50, 2.57) |
| Adjusted* | REF | 0.60 (0.23, 1.53) | REF | 1.00 (0.42, 2.43) |
| Adjusted* CN<br>only** | REF | 0.49 (0.19, 1.32) | REF | 1.13 (0.46, 2.80) |

\*Adjusted for study, age, sex, BMI, and brain atrophy
\*\*n=186
CI=confidence interval, CN=cognitively normal, OR=odds ratio, REF=reference

## Discussion

In this analysis of community dwelling older adults, we found that participants with higher dopaminergic integrity in the sensorimotor striatum and associative striatum were more likely to maintain their gait speed during dual-task walking without increased PFC effort than to do so with increased PFC activation. In addition, greater dopaminergic integrity in the sensorimotor striatum only was related to lower likelihood of being in the group that declined performance without engagement of additional PFC effort compared to the group that maintained gait performance without increasing PFC effort, though these findings were less robust. The highest DTBZ binding was consistently found in the group that maintained gait performance without increasing PFC effort (PFC-/gait+). This is presumably the group with the most automatic control of gait^3^ who can effectively adapt to the demands of dual-task walking without additional engagement of the PFC. Contrary to our hypothesis, there were no statistical differences in dopaminergic integrity for either the sensorimotor or associative region with being in the group that declined in gait performance despite increased PFC effort. Notably, no differences were observed when groupings were made using either PFC activation alone or gait performance alone, suggesting that neither component alone is driving the observed associations.

Prior research has suggested that polymorphisms of the catecholamine-O-methyl-transferase (COMT) gene related to greater dopaminergic signaling in the PFC are related to higher gait speed both cross-sectionally and longitudinally^35-37^. In addition, older adults with mild parkinsonian signs including slow gait speed had lower functional PFC-striatal connectivity^38^. These prior results suggest that dopaminergic integrity in older adults may play a critical role in how the PFC is involved in control of mobility.

However, there is limited data assessing how nigrostriatal dopaminergic integrity is related to mobility and motor control in older adults without Parkinson’s disease and no prior studies of dopamine and PFC-mediated motor control during walking.

In our analyses, DTBZ binding of both regions was lower in the group that had effortful maintenance of gait performance (PFC+/gait+) compared to those who maintained gait performance without extra PFC resources (PFC-/gait+) which may signify reduced automaticity and increased need for PFC engagement to maintain performance. We hypothesized that this group would have lower dopaminergic integrity of the sensorimotor region but higher dopaminergic integrity of the associative region compared to the PFC-/gait+ group, which would facilitate the compensatory signaling in the PFC. While this compensatory engagement of the PFC during walking is important for maintenance of performance under challenging conditions such as dual-tasking, it may come at the expense of availability of PFC resources for dealing with other environmental threats that could lead to falls^5^. Of note, this group may also be at greater risk for falls based on their lower dopamine levels^39^, potentially compounding the risk from PFC inefficiencies.

We had hypothesized that the groups that declined in gait speed from single-to dual-task would be the groups with the lowest dopaminergic binding in both regions. However, this was only observed for the group that declined in performance with no additional activation of the PFC (PFC-/gait-) and only for the sensorimotor striatum, suggesting a loss of automaticity in this group without compensatory PFC signaling, possibly due to neural pathologies that we did not include here. We observed no differences in DTBZ binding for those in the PFC+/gait-group compared to the PFC-/gait+ group. It may be that the effects of loss of dopaminergic integrity differ across individuals based on the health of other neurologic systems, facilitating maintenance of dopaminergic neurotransmission and preservation of function in the presence of presynaptic losses in nigrostriatal dopamine neurons. This is supported by the fact that VMAT2 reductions may be >50% before clinical Parkinson’s disease is detected. We assessed some other aspects of neurologic health (volume of WMH^40^ and brain atrophy) as confounders in our analyses but there are other potential aspects that could interact with dopaminergic function, such as cholinergic function^41,42^ and functional connectivity^38,43^, to determine performance.

There are several limitations of this study to note. These analyses were cross-sectional so we are unable to determine directionality of these associations. Our sample had limited generalizability and primarily represents White individuals with higher educational attainment. Our interpretation of DTBZ specific binding outcomes in sensorimotor and associative striatum as an index of the density of presynaptic terminals of the nigrostriatal dopamine projection is predicated on previous preclinical evidence that VMAT2 expression is linearly related to the integrity of nigrostriatal dopamine neurons and that VMAT2 expression is insensitive to compensatory regulation attributed to lesion or dopamine-enhancing drugs^29,44^. Finally, we had a limited sample size within our PFC/gait groups, which may have limited our power to detect true differences in some cases. However, this is the first sample to our knowledge to assess both PFC activation during complex walking tasks using fNIRS and dopaminergic neurotransmission in the striatum using PET. In addition, we considered the combination of fNIRS signal and gait performance in order to disentangle adaptive vs non-adaptive PFC activation.

In conclusion, we found that higher dopaminergic integrity in both the sensorimotor striatum and the associative striatum was related to a lower likelihood of maintaining gait performance during dual-task walking with increased PFC effort than those who did so without increasing PFC effort. Dopaminergic therapy may prove useful for older adults without Parkinson’s disease for maintenance of gait automaticity, though a better understanding of how higher dopaminergic neurotransmission is related to compensatory compared to inefficient PFC signaling is needed.

**Supplementary Table 1.** Demographic, health, neuroimaging, and dual task walking characteristics by study.

|  | **MMH** | **SOMMA** | **Total** |
| --- | --- | --- | --- |
|  | N=85 | N=116 | N=201 |
|  | Mean (SD) or n (%) | Mean (SD) or n (%) | Mean (SD) or n (%) |
| Age (years) | 72.3 (4.3) | 76.7 (3.9) | 74.9 (4.6) |
| Women | 57 (67.1%) | 70 (60.3%) | 127 (63.2%) |
| White race | 78 (91.8%) | 107 (92.2%) | 185 (92.0%) |
| > HS education | 59 (69.4%) | 106 (92.2%) | 165 (82.5%) |
| BMI | 30.9 (6.0) | 26.6 (4.3) | 28.4 (5.5) |
| Systolic blood pressure (n=173) | 132.4 (16.9) | 129.9 (14.0) | 130.8 (15.0) |
| Any health condition | 44 (52.4%) | 56 (48.3%) | 100 (50.0%) |
| Cognitively unimpaired | 80 (94.1) | 106 (92.2%) | 186 (93.0%) |
| WMH (%; n=197) | 0.012 (0.029) | 0.016 (0.022) | 0.014 (0.02) |
| Brain atrophy | 0.35 (0.03) | 0.33 (0.03) | 0.34 (0.03) |
| *DTBZ binding* |  |  |  |
| Sensorimotor striatum | 2.46 (0.41) | 2.43 (0.45) | 2.44 (0.43) |
| Associative Striatum | 2.09 (0.32) | 2.09 (0.36) | 2.09 (0.34) |
| *Dual-Task* |  |  |  |
| HbO, dual task-single task (t-stat) | 0.72 (2.28) | 0.83 (2.16) | 0.78 (2.21) |
| Gait speed change dual task-single task (m/s) | -0.11 (0.09) | -0.14 (0.14) | -0.13 (0.12) |
| Gait speed single task (m/s) | 1.06 (0.18) | 1.13 (0.21) | 1.10 (0.20) |
| Rate of correct letters dual task (letters/s) | 0.58 (0.16) | 0.60 (0.20) | 0.60 (0.19) |

HS=high school, BMI=body mass index, WMH=white matter hyperintensities, DTBZ=dihydrotrabenazine, HbO=oxygenated hemoglobin, m/s=meters/second

## Data Availability

All data used in this study are available upon request from the parent studies which includes the Study of Muscle, Mobility and Aging (SOMMA) and the Mon-Yough Healthy Aging Team (MYHAT)

https://www.sommastudy.com/

https://www.dementia-epidemiology.pitt.edu/myhat/

## Notes

**Funding statement:** The MYHAT study was supported by the National Institute on Aging (R01AG023651). The Study of Muscle, Mobility and Aging is supported by funding from the National Institute on Aging (R01 AG 059416). Study infrastructure support was funded in part by NIA Claude D. Pepper Older American Independence Centers, at University of Pittsburgh (P30 AG024827) and Wake Forest University (P30 AG021332) and the Clinical and Translational Science Institutes, funded by the National Center for Advancing Translational Science, at Wake Forest University (UL1 TR001420). This analysis was supported by NIA awards (R01AG075025 and U01AG061393).

### Competing Interest Statement

The authors have declared no competing interest.

### Funding Statement

The MYHAT study was supported by the National Institute on Aging (R01AG023651). The Study of Muscle, Mobility and Aging is supported by funding from the National Institute on Aging (R01 AG 059416). Study infrastructure support was funded in part by NIA Claude D. Pepper Older American Independence Centers, at University of Pittsburgh (P30 AG024827) and Wake Forest University (P30 AG021332) and the Clinical and Translational Science Institutes, funded by the National Center for Advancing Translational Science, at Wake Forest University (UL1 TR001420). This analysis was supported by NIA awards (R01AG075025 and U01AG061393).

### Author Declarations

All data are available from the parent studies which includes the Study of Muscle, Mobility and Aging (SOMMA, https://www.sommastudy.com/) and the Mon-Yough Healthy Aging Team (MYHAT, https://www.dementia-epidemiology.pitt.edu/myhat/)

